# A rapid review of Supplementary air filtration systems in health service settings. September 2022

**DOI:** 10.1101/2022.10.25.22281493

**Authors:** Charlotte Bowles, Tom Winfield, Lauren Elston, Elise Hasler, Antonia Needham, Alison Cooper, Ruth Lewis, Adrian Edwards

## Abstract

The aerosol spread of SARS-CoV-2 has been a major challenge for healthcare facilities and there has been increased use of supplementary air filtration to mitigate SARS-CoV-2 transmission. Appropriately sized supplementary room air filtration systems could greatly reduce aerosol levels throughout ward spaces. Portable air filtration systems, such as those combining high efficiency particulate air (HEPA) filters and ultraviolet (UVC) light sterilisation, may be a scalable solution for removing respiratory viruses such as SARS-CoV-2. This rapid review aimed to assess the effectiveness of supplementary air cleaning devices in health service settings such as hospitals and dental clinics (including, but not limited to HEPA filtration, UVC light and mobile UVC light devices) to reduce the transmission of SARS-CoV-2.

One systematic review (Daga et al. 2021), three observational studies (Conway Morris et al. 2022, Thuresson et al. 2022, Sloof et al. 2022), one modelling study, (Buchan et al. 2020) and two experimental studies (Barnewall & Bischoff 2021, Snelling et al. 2022) were found. Outcome measures included symptom scores, presence of SARS-CoV-2 RNA in sample counts, general particulate matter counts, viral counts, and relative risk of SARS-CoV-2 exposure. From real world settings, the systematic review assessed the effectiveness of HEPA filtration in dental clinics (Daga et al. 2021), two additional observational studies assessed HEPA and UV light in UK hospital settings (Conway Morris et al. 2022, Sloof et al. 2022) and one observational study included mobile HEPA-filtration units in Swedish hospitals (Thuresson et al. 2022). Studies were published from 2020 onwards.

Real world evidence suggests supplementary air systems have the potential to reduce SARS-CoV-2 in the air and subsequently reduce transmission or infection rates but further research, with study designs having lower risk of bias, is required. HEPA filters alongside UVC light could provide the most notable reductions in SARS-CoV-2 counts, although the supporting evidence relates to HEPA/UVC filtration, and this review does not provide evidence on the effectiveness of other potential supplementary air filtration systems that could be used. Evidence is limited on the optimum air changes per hour needed and the positioning of air filtration units in rooms.

**Funding statement:** The Wales Centre for Evidence Based Care was funded for this work by the Wales COVID-19 Evidence Centre, itself funded by Health & Care Research Wales on behalf of Welsh Government.

---

**Wales COVID-19 Evidence Centre (WCEC) Rapid Review**

**Supplementary air filtration systems in health service settings Report number – RR00041 September 2022**

**Rapid Review Details**

**Review conducted by:** Health Technology Wales (HTW)

**Review Team:**

▪ Charlotte Bowles, Health Technology Wales,
▪ Tom Winfield, Health Technology Wales,
▪ Lauren Elston, Health Technology Wales,
▪ Elise Hasler, Health Technology Wales,
▪ Antonia Needham, Health Technology Wales

**Review submitted to the WCEC on:**

20^th^ September 2022

**Stakeholder consultation meeting:**

21^st^ September 2022

**Rapid Review report issued by the WCEC on:**

October 2022

**WCEC Team:**

▪ Adrian Edwards, Alison Cooper, Ruth Lewis, Micaela Gal and Jane Greenwell involved in drafting Topline Summary and editing

**This review should be cited as:**

RR00041.Wales COVID-19 Evidence Centre. A rapid review looking at supplementary air filtration systems in health service settings. October 2022

**This report can be downloaded here:**

https://healthandcareresearchwales.org/wales-covid-19-evidence-centre-report-library

**Disclaimer:** The views expressed in this publication are those of the authors, not necessarily Health and Care Research Wales. The WCEC and authors of this work declare that they have no conflict of interest.

**FULL REPORT**

**TOPLINE SUMMARY**

**What is a Rapid Review?**

Our rapid reviews use a variation of the systematic review approach, abbreviating or omitting some components to generate the evidence to inform stakeholders promptly whilst maintaining attention to bias. They follow the methodological recommendations and minimum standards for conducting and reporting rapid reviews, including a structured protocol, systematic search, screening, data extraction, critical appraisal, and evidence synthesis to answer a specific question and identify key research gaps. They take 1-2 months, depending on the breadth and complexity of the research topic/ question(s), extent of the evidence base, and type of analysis required for synthesis.

**Who is this summary for?**

The Science Evidence Advice in the Welsh Government.

**Background / Aim of Rapid Review**

The aerosol spread of SARS-CoV-2 has been a major challenge for healthcare facilities and there has been increased use of supplementary filtration to mitigate SARS-CoV-2 transmission. Appropriately sized supplementary room air filtration systems could greatly reduce aerosol levels throughout ward spaces. Portable air filtration systems, such as those combining high efficiency particulate air (HEPA) filters and ultraviolet (UVC) light sterilisation, may be a scalable solution for removing respiratory viruses such as SARS-CoV-2. This rapid review aimed to assess the effectiveness of supplementary air cleaning devices in health service settings such as hospitals and dental clinics (including, but not limited to HEPA filtration, UVC light and mobile UVC light devices) to reduce the transmission of SARS-CoV-2.

**Key Findings**

*Extent of the evidence base*

▪ We found one systematic review (Daga et al. 2021), three observational studies (Conway Morris et al. 2022, Thuresson et al. 2022, Sloof et al. 2022), one modelling study, (Buchan et al. 2020) and two experimental studies (Barnewall & Bischoff 2021, Snelling et al. 2022).
▪ Outcome measures included symptom scores, presence of SARS-CoV-2 RNA in sample counts, general particulate matter counts, viral counts, and relative risk of SARS-CoV-2 exposure.
▪ From real world settings, the systematic review assessed the effectiveness of HEPA filtration in dental clinics (Daga et al. 2021), two additional observational studies assessed HEPA and UV light in UK hospital settings (Conway Morris et al. 2022, Sloof et al. 2022) and one observational study included mobile HEPA-filtration units in Swedish hospitals (Thuresson et al. 2022)

*Recency of the evidence base*

▪ Studies were published from 2020 onwards.

*Evidence of effectiveness*

▪ There is some evidence on the effectiveness of supplementary air filtration systems in healthcare settings to reduce SARS-CoV-2 transmission although some studies have high risk of bias.
▪ UVC light and/or HEPA filtration can significantly reduce particulate matter counts and may also lead to lower symptom scores of COVID-19.
▪ Two trials in the systematic review reported lower COVID-19 symptom scores with HEPA filtration (Daga et al. 2021).
▪ Thuresson et al. (2022) found that adding a mobile HEPA-filtration unit to rooms with regular ventilation was associated reduced SARS-CoV-2 RNA in the air.
▪ HEPA/UV filtration units greatly reduced particulate matter counts of all sizes (Sloof et al. (2022).
▪ Similar reductions in particulate matter counts were reported in the modelling study: Far-UVC light (compared to no UVC light) greatly reduced viral counts, and when combined with eight air changes per hour (ACH), viral removal was much quicker when compared with 0.8 ACH.
▪ Barnewall & Bischoff (2021) found particulate matter counts reduced most when UVC light and a HEPA filter were used together.
▪ Snelling et al. (2022) found that UVC combined with five ACH completely inactivated Bovine CoV bioaerosols (a surrogate for SARS-CoV-2).

*Critical Appraisal*

▪ Two studies, (Conway Morris et al. 2022, Daga et al. 2021) were assessed as having high risk of bias concerns.
▪ Two observational studies (Sloof et al. 2022, Thuresson et al. 2022) and the modelling study (Buchan et al. 2020) were assessed as having low risk of bias.

**Policy Implications**

▪ Real world evidence suggests supplementary air systems have the potential to reduce SARS-CoV-2 in the air and subsequently reduce transmission or infection rates but further research, with study designs having lower risk of bias, is required.
▪ HEPA filters alongside UVC light could provide the most notable reductions in SARS-CoV-2 counts, although the supporting evidence relates to HEPA/UVC filtration, and this review does not provide evidence on the effectiveness of other potential supplementary air filtration systems that could be used.
▪ Evidence is limited on the optimum air changes per hour needed and the positioning of air filtration units in rooms.

**Strength of Evidence**

▪ This review is limited to a few studies conducted within real world healthcare settings.
▪ We are reliant on interpreting the results of studies that have several limitations and this reduces the strength of conclusions.

## 1. BACKGROUND

### 1.1 Who is this review for?

This Rapid Review is being conducted as part of the Wales COVID-19 Evidence Centre Work Programme. The original research topic area was submitted through the Cwm Taf Morgannwg Innovation Portal. The research question was developed through collaboration with a range of stakeholders including from the COVID-19 Technical Advisory Cell (Welsh Government), the WCEC Core Team, and Health Technology Wales.

### 1.2 Purpose of this review

During the COVID-19 pandemic, the aerosol spread of SARS-CoV-2 in healthcare settings has been a major problem, resulting in increased use of supplementary air filtration systems to mitigate transmission of SARS-CoV-2 (Sloof et al. 2022). Despite the use of personal protective equipment during the pandemic, there is a need to improve the safety for healthcare workers and patients by finding other ways to decrease the airborne transmission of SARS-CoV-2 and other viruses. Therefore, the purpose of this review is to evaluate the effectiveness of supplementary air cleaning devices specifically in healthcare settings.

Established technologies such as air filtration, and ultraviolet-C (UVC) light have the potential to reduce the transmission of airborne viruses. Supplementary air filtration systems vary and can include, but are not limited to, UVC irradiation and standalone HEPA filtration units. Portable air filtration systems, which combine high efficiency particulate filtration and ultraviolet (UV) light sterilization, may also be a solution for removing respirable SARS-CoV-2 and other viruses (Conway Morris et al. 2022). Appropriately sized supplementary room air filtration systems, if utilised correctly, can greatly reduce rates of nosocomial viral infection in such settings and warrants further investigation (Sloof et al. 2022). Sloof et al. (2022) highlights the importance of commissioning such devices, considering their effect on air flow and the removal of contaminants to optimise the system’s ability to clean the air in the ward space.

Ultraviolet has a well-known antiviral effect (Heßling et al. 2020) and the use of UVC irradiation as a means of microbial inactivation is well established in multiple sectors including medical, scientific, manufacturing, and agricultural settings (C Hopkins 2022, personal communication, 28 September). UVC is germicidal at certain wavelengths of the light spectrum by damaging microorganisms rendering them unable to replicate. Ultraviolet germicidal irradiation (UVCI) is defined as the use of ultraviolet (UV) wavelengths of light in the germicidal rang of 200-320nm for the disinfection of air and surfaces (Kowalski 2009).

The term UVC lamp specifically refers to lamps that produce UVC wavelengths in the narrow band range of 200-280nm (Kowalski 2009). Low pressure mercury lamps produce a narrow band of UVC at about 254nm (Kowalski 2009, Heßling et al. 2020). This is known as Germicidal Ultraviolet (GUV) or Ultraviolet Germicidal Irradiation (UVGI). UVGI has previously been considered as a way of controlling airborne viruses during a pandemic if effective vaccines or antiviral drugs are not available (Buchan et al. 2020).

Ozone generation is identified among the risks associated with UV disinfection, especially for air disinfection application (Raeiszadeh & Adeli 2020). Ozone, an allotrope of oxygen, can be produced when oxygen is exposed to UVC with a wavelength below 240nm. Ozone above occupational exposure limits is harmful to human health and can affect the respiratory, cardiovascular, and central nervous system (C Hopkins 2022, personal communication, 28 September). Ozone can also cause degradation of certain materials, which can lead to fire hazards. As a result of this, regulations are required to prevent the production of ozone by UVC systems (C Hopkins 2022, personal communication, 28 September).

### 1.3 Research Question

This rapid review aims to provide evidence on the following research question:

□ What is the effectiveness of supplementary air cleaning devices in health service settings to reduce the transmission of SARS-CoV-2?

We searched for evidence on the effectiveness of supplementary air cleaning devices in health service settings such as hospitals and dental clinics (including, but not limited to HEPA filtration, UVC light and mobile UVC light devices) to reduce the transmission of SARS-CoV-

More detail on the study selection methods and study inclusion criteria is provided in Section 5.

## 2. FINDINGS

### 2.1 Overview of the available evidence

We searched for published articles where the effectiveness of any supplementary air filtration systems was reported. As per the protocol, we prioritised studies conducted in real world settings (i.e., hospitals or other healthcare settings such as dental clinics). Where data was not available, we also included evidence from modelling studies that aligned with the research question and eligibility criteria. Additionally, we have reported in outcomes for other pathogens where the paper reported evidence relating to SARS-CoV-2, as stated in the protocol (see methods). Our search generated 6,201 references that were screened against the eligibility criteria (see methods). In total, seven studies met the inclusion criteria and were included in this review (see Tables 2-4). We only included studies that were related to healthcare settings to align with the study inclusion criteria, which led to the exclusion of some studies that reported the effectiveness of supplementary air filtration systems in other settings. Appendix 3 details a list of additional studies that were identified by stakeholders, with reasons for exclusion.

**Table 1:**
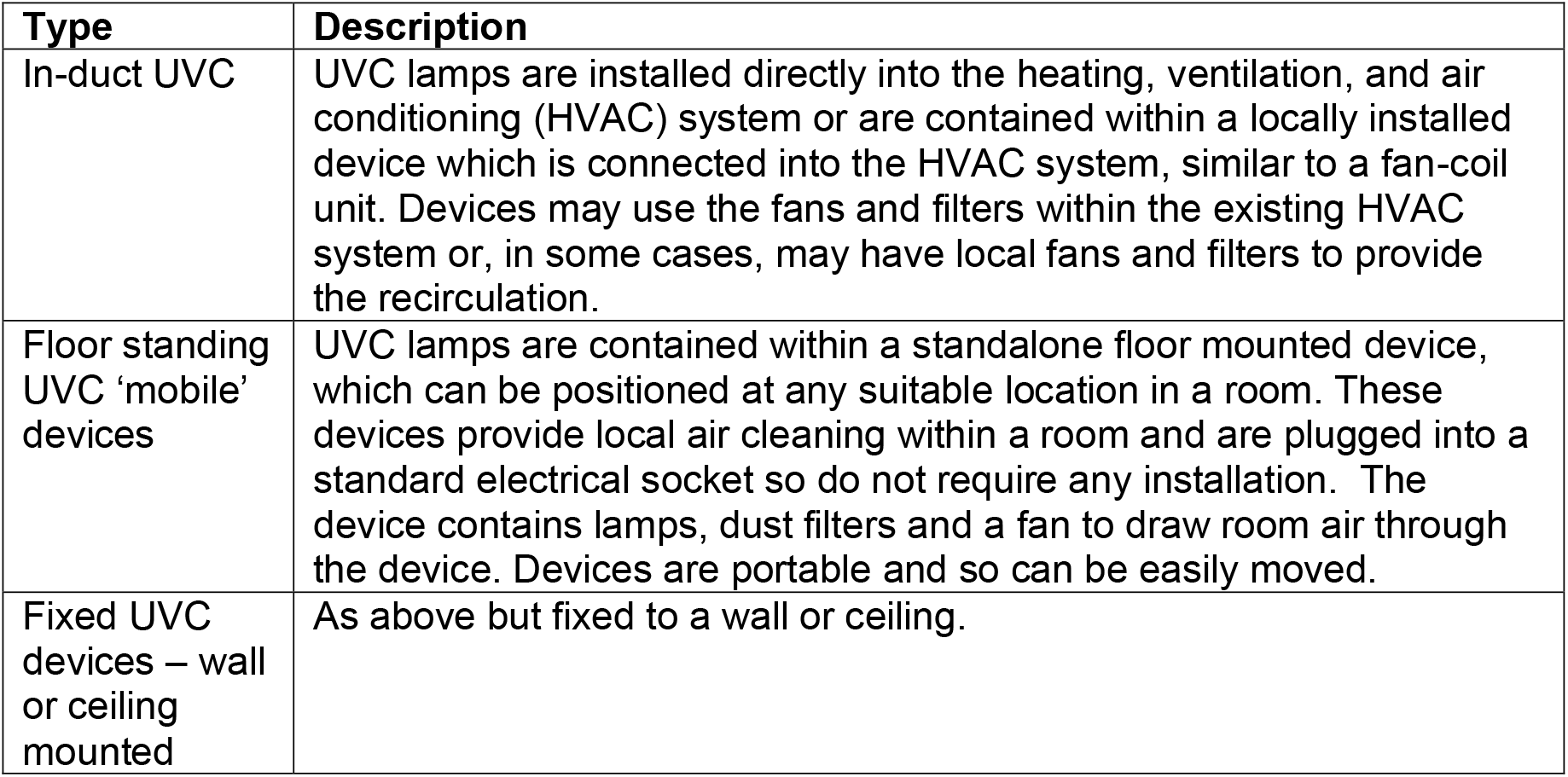
Types of 254nm UVC devices (C Hopkins 2022, personal communication, 28 September)

**Table 2:**
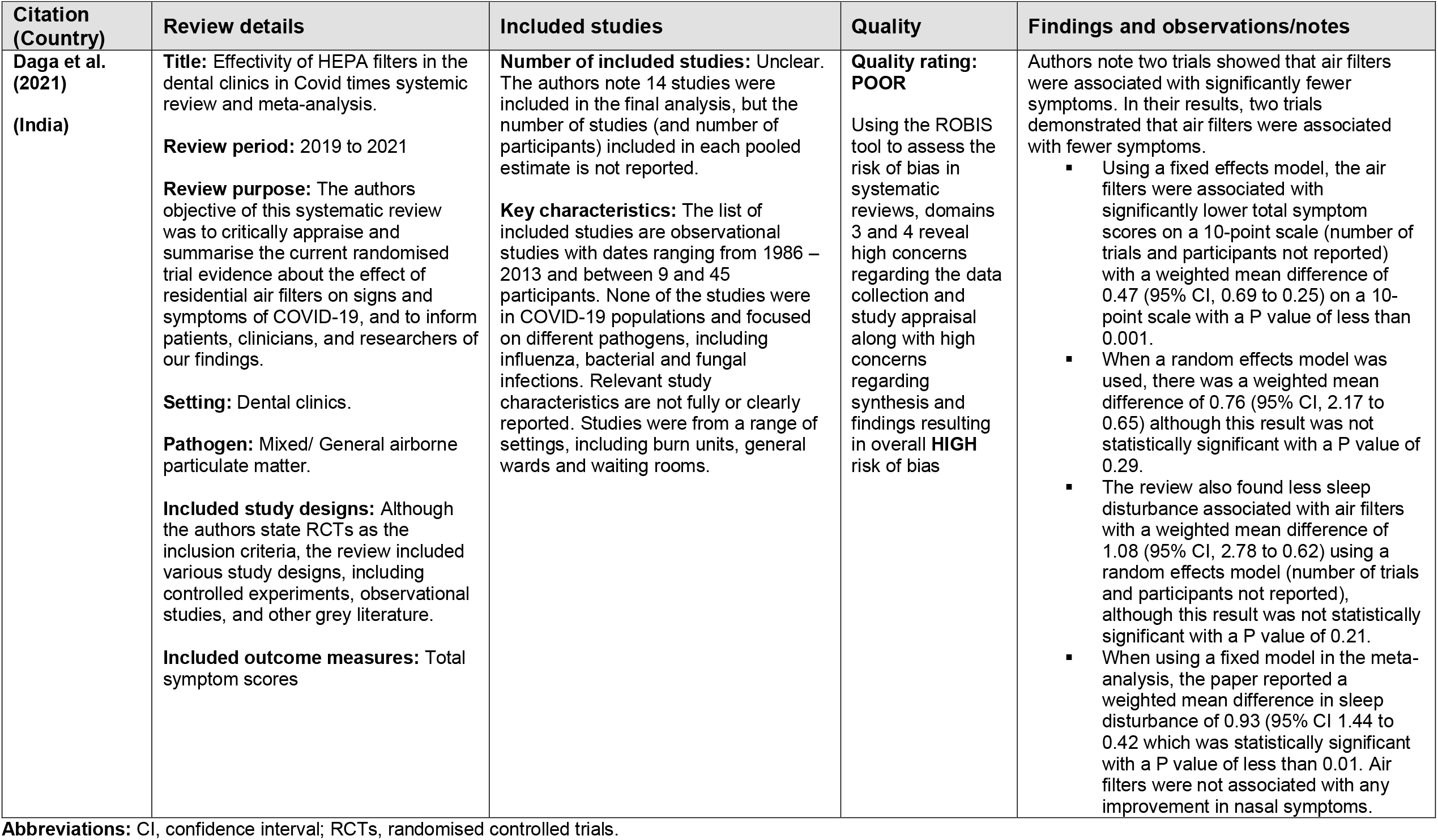
Summary of Systematic Reviews.

**Table 3:** Summary of Primary Studies.

| Citation (Country) | Study Details | Participants & setting | Key findings |
| --- | --- | --- | --- |
| **Conway Morris et al. (2022)**<br>**(UK)** | **Title:** The removal of airborne severe acute respiratory syndrome coronavirus 2 (SARS-CoV-2) and other microbial bioaerosols by air filtration on coronavirus disease 2019 (COVID-19) surge units.<br>**Study Design:** Prospective experimental cohort study<br>**Type of intervention [exposure]:** AC1500 HEPA14/UVC sterilizer (Filtrex, Harlow, UK); in the ICU: a Medi 10 HEPA13/UVC sterilizer (Max Vac, Zurich, Switzerland). The air filters were placed in fixed positions before the initiation of the three-week study period.<br>**Aims:** The study aimed to provide evidence for the removal of SARS-CoV-2 and microbial bioaerosols from the air using portable air filters with UVC sterilization on a COVID-19 ward.<br>**Data collection methods:** Crossover evaluation, with the primary outcome being detection of SARS-CoV-2 RNA in the various size fractions of air samples. The air filters were placed before the initiation of the three-week study period and switched on at the beginning of week 2 and run continuously from Sunday to Sunday for 24 hours per day. Air sampling was conducted using National Institute for Occupational Safety and Health (NIOSH) BC 251 2-stage cyclone aerosol samplers. Samplers were operated on weekdays (from 08:15 to 14:15) for 3 consecutive weeks.<br>**Quality rating: POOR** | **Pathogen:** SARS-CoV-2<br>**Setting:** 2 repurposed COVID-19 units. One surge ward (ward) and one surge intensive care ward (ICU).<br>**Dates of data collection:** 18 January to 5 February 2021 | A crossover evaluation was performed where samples were taken to detect SARS-CoV-2 RNA before and after the HEPA/UVC steriliser air filter was introduced in the ward and unit.<br>**Primary Findings:** The study found that after the portable air filter was running for two full weeks in the ward, SARS-CoV-2 RNA was not detected in any of the sampling fractions on any of the 5 testing days; whereas during the first week while the air filter was inactive, SARS-CoV-2 was detected on all sampling days; RNA was detected in both the medium (1–4 micrometres ($\mu\text{M}$)) and the large ($>4 \mu\text{M}$) particulate fractions. SARSCoV-2 RNA was not detected in the small ($<1 \mu\text{M}$) particulate filter.<br>**Additional Findings:** In the ICU, there was limited evidence of airborne SARS-CoV-2 in weeks 1 and 3 (filter off) but detected SARS-CoV-2 RNA in a single sample in the medium (1–4 $\mu\text{M}$) particulates on week 2 (filter on). |
| **Thuresson et al. (2022)**<br>**(Sweden)** | **Title:** Airborne severe acute respiratory syndrome coronavirus 2 (SARS-CoV-2) in hospitals: effects of aerosol-generating procedures, HEPA-filtration units, patient viral load, and physical distance.<br>**Study design:** Prospective exploratory observational study<br>**Type of intervention:** Different ventilation and patient characteristics were investigated, and this included mobile HEPA-filtration units.<br>**Aims:** To identify factors associated with airborne SARS-CoV-2 RNA, including potential aerosol generating procedures, patient characteristics, and environmental parameters. | **Pathogen:** SARS-CoV-2<br>**Setting:** Two hospitals in Sweden, including 3 infection disease wards, 4 ICUs, 3 medical wards converted to COVID-19 units, and 1 emergency department. | **Primary Findings:** Compared with rooms with normal ventilation and no HEPA unit ($n = 57$ samples), rooms with normal ventilation plus an additional mobile HEPA-filtration unit ($n = 139$ samples) were associated with a reduction in SARS-CoV-2 RNA in the air (OR 0.3, 95% CI 0.12 to 0.98, $p = 0.02$).<br>Compared with rooms with normal ventilation (3-4 ACH), enhanced ventilation of 8 ACH was associated with a reduction in SARS-CoV-2 RNA in the air (OR 0.3, 95% CI 0.13 – 0.96, $p = 0.02$). |
|  | **Data collection methods:** Data was collected from several wards treating patients with COVID-19 across 2 hospitals. Ventilation in each room was a nominal 3–4 air ACH. Six rooms at one of the infectious disease wards had an installed higher ventilation rate of 8 ACH. As an addition to regular ventilation, mobile HEPA filtration units, delivering approximately 200L filtered air per second, were used at the discretion of the ward staff, but were recommended when aerosol generating procedures were performed. Rooms with a mobile HEPA-filtration unit or a high ventilation rate (8–9 ACH). Most rooms were single or double rooms, but the ICUs also had larger cohort rooms for up to 16 patients and used up to 4 mobile HEPA-filtration units. Samples were collected using a liquid cyclone (Coriolis µ; Bertin Instruments, France), and sampling in patient rooms was performed at a predefined distance of less than 1 m, 1–2 m, or 2–4 m from the patient's head.<br>**Quality rating: GOOD** | **Dates of data collection:** March 2020 to April 2021 |  |
| **Sloof et al. (2022)**<br>**(UK)** | **Title:** Impact of supplementary air filtration on PM in a UK hospital ward.<br>**Study Design:** pre-print prospective observational study.<br>**Type of intervention [exposure]:** AFU on an inpatient ward containing HEPA filters and an UVC air disinfection lamp (at 254 nm)<br>**Aims:** The study aimed to assess aerosol transport within the ward and determine whether an AFU reduced PM levels and CO<sub>2</sub> levels in the air.<br>**Data collection methods:** Time-series PM, CO<sub>2</sub>, temperature, and humidity data (at 1-minute intervals) was collected from multiple sensors around the ward over two days in August 2021. During this period, the AFU was accidentally switched off for approximately 7 hours, allowing the impact of the intervention on particulates (PM1-PM10) to be assessed using a Mann-Whitney test. Pearson correlation analysis of the PM and CO<sub>2</sub> signals was also undertaken to evaluate the movement of airborne particulates around the ward.<br>**Quality rating: GOOD** | **Pathogen:** General airborne PM.<br>**Setting:** Hospital ward.<br>**Dates of data collection:** 3 and 4 August 2021 | **Primary Findings:** The data was collected from seven automatic sensors every one minute, giving a total of 2782 data points per sensor over the two-day period. However, actual baseline ventilation rates that occurred on the 3rd and 4th August are not reported in the paper as they were unknown.<br>The statistical analysis found that for all the sensors the action of the AFU was associated with a large effect on PM (PM1, PM2.5, CO<sub>2</sub> and VP levels, which was highly significant.<br>The primary results of the paper found that the AFU greatly reduced PM counts of all sizes throughout the ward space, with a P value of less than 0.001 for all sensors. The PM signals positively correlated with indoor CO<sub>2</sub> levels ($r = 0.343 - 0.817$; with all values reported as statistically significant. |

**Abbreviations:** AFU, air filtration unit; HEPA, high efficiency particulate air; VP, vapour pressure; PM, particulate matter; ACH, air changes per hour.

**Table 4:** Summary of Modelling and Experimental Studies.

| Study | Title/Purpose | Analysis | Intervention | Control/comparison | Outcomes |
| --- | --- | --- | --- | --- | --- |
| Buchan et al. (2020)<br>High-fidelity coupled radiation-CFD model<br>(UK/Canada)<br>ROB concern: **LOW** | **Title:** Predicting airborne coronavirus inactivation by far-UVC in populated rooms using a high-fidelity coupled radiation-CFD model.<br>**Purpose:** A modelling study, presenting a coupled radiation transport and fluid dynamics simulator, based on the Boltzmann Transport and Navier–Stokes equations with integrated Large Eddy Simulation (LES) turbulence models, for viral inactivation within atmospheres. | **Model:**<br>Far-UVC radiation transport model.<br>Fluid flow model for room ventilation.<br>UVC inactivation model. | Quantifying the rate of far-UVC viral inactivation. | With and without far-UVC (room's ventilation alone)<br>8.0 ACH versus 0.8 ACH | Total viral concentration over time: Without the lamp, 0.8 air changes per hour (ACH) ventilation results in very slow reductions, but when increased to 8.0 ACH, viral removal through ventilation begins 45 s after release and concentrations are reduced by 90% and 99% in approximately 12 and 24 minutes, respectively.<br>Viral count: The combination of far-UVC and high ventilation reduces the viral count most effectively, times to achieve 90% and 99% reductions being approximately 6 and 11.5 minutes, respectively, more than halving the times when using 8.0 ACH ventilation alone. |
| Barnewall & Bischoff (2021)<br>Research brief paper<br>(US) | **Title:** Removal of SARS-CoV-2 bioaerosols using ultraviolet air filtration.<br>**Purpose:** An experimental study, testing the efficacy of UVC light and HEPA filtration with an air purification system (APS) to remove SARS-CoV-PM counts. | **Methods:** The APS consisted of a UVC photolytic chamber, which incorporated 4 low-pressure UVC germicidal lamps operating at the 254-nm wavelength at a total irradiance of 177.8 $\mu\text{W}/\text{cm}^2$ and containing ~2,400 transparent quartz tubular elements oriented randomly in the chamber. | Air purification system (APS) combining UVC light and high-efficiency particulate air (HEPA) filtration in a controlled environment using SARS-CoV-2 as test organism. | 3 test groups:<br>Inactive test unit (control)<br>APS with UVC light<br>APS with UVC light and HEPA filter. | In the control group, particle counts increased from 20 (in the pre-APS chamber) to 54 particles (in the post-APS chamber) after the introduction of the SARS-CoV-2 aerosol.<br>In the APS with UVC light, particle counts increased from 79 in the pre-APS chamber to 101 in the post-APS chamber.<br>Placing a HEPA filter into the airflow reduced the particle count from 79 to 5 particles. |
| Snelling et al. (2022)<br>Scenario-based modelling<br>(UK/Northern Ireland) | **Title:** Efficacy of a single pass UVC air treatment for the inactivation of coronavirus, MS2 coliphage and Staphylococcus aureus bioaerosols.<br>**Purpose:** To investigate the efficacy of a high velocity, single pass system to deactivate relevant model pathogens (MS2, Staphylococcus aureus, bovine coronavirus) | **Methods:** Scenario-based modelling was used to investigate the reduction in risk of airborne person-to-person transmission based upon a single infected subject within the small room. | UVC air treatment device with HEPA filter. Three consecutive UVC exposure chambers. | Assumed no comparison/N.R. | Bovine CoV was used as a surrogate for SARS-CoV2<br>Single pass UVC air treatment (1254 L/min) can effectively inactivate MS2 (6-log reduction), S. aureus (4.7-log reduction) and bovine coronavirus (2.4-log reduction) |

**Abbreviations:** CFD, Computational Fluid Dynamics; UVC, ultraviolet C; AFU, air filtration unit; HEPA, high efficiency particulate air; PM, particulate matter; ACH, air changes per hour

We identified one systematic review (Daga et al. 2021), three primary studies, (Conway Morris et al. 2022, Thuresson et al. 2022, Sloof et al. 2022) one modelling study (Buchan et al. 2020) and two experimental studies (Barnewall & Bischoff 2021, Snelling et al. 2022). The purpose of each study varied in the type of air filtration unit used along with different settings across studies. The systematic review, Daga et al. (2021) explored the effectivity of HEPA filters in the dental clinics, whereas the primary studies identified in this rapid review looked at HEPA/UVC sterilisers/filters in hospital settings. The study aims, objectives, outcome measures and methods differed across the modelling and experimental studies. One modelling study (Buchan et al. 2020) looked at the efficacy of UVC, and two experimental studies, (Barnewall & Bischoff 2021, Snelling et al. 2022) tested the efficacy of UVC light with HEPA filters.

### 2.2 Summary of the evidence

#### 2.2.1 Systematic review

One systematic review and meta-analyses (Daga et al. 2021) reported on the effectivity of HEPA filters in healthcare settings. Although the focus within the review was on COVID-19 populations and symptoms, the included evidence was pre-pandemic and evaluated HEPA filters with other pathogens. The review included 14 studies, although the reported number of studies, and design of studies, were inconsistently reported throughout the review. The overall risk of bias report for this review revealed high concerns, and it is also unclear from the review how many studies (or which studies) were included in each of the pooled outcomes of the meta-analysis, nor which studies relate to the outcomes described in the narrative synthesis. Furthermore, the authors synthesised symptom scores on a 10-point scale, but do not report what scale is used, or whether all studies used the same symptom scale; the authors did note that none of the studies used a validated scale. Outcomes relating to the presence and transmission of the SARS-CoV-2 were not reported in the paper.

In their results, two trials demonstrated that air filters were associated with fewer symptoms than when no filters were used. Using a fixed effects model, the air filters were associated with significantly lower total symptom scores with a weighted mean difference of 0.47 (95% confidence interval [CI], 0.69 to 0.25) on a 10-point scale with a P value of less than 0.001 (Daga et al. 2021). However, when a random effects model was used, there was a weighted mean difference of 0.76 (95% CI, 2.17 to 0.65) but this result was not statistically significant, with a P value of 0.29 (Daga et al. 2021).

The review also found less sleep disturbance associated when air filters were used when compared with no filters used, with a weighted mean difference of 1.08 (95% CI, 2.78 to 0.62) using a random effects model, although this result was not statistically significant with a P value of 0.21 (Daga et al. 2021). For sleep disturbance, when using a fixed model in the meta-analysis, the paper reported a weighted mean difference of 0.93 (95% CI 1.44 to 0.42 which was statistically significant with a P value of less than 0.01 (Daga et al. 2021). The heterogeneity values for this review were not reported, although the paper stated that the heterogeneity of results weakens the inferences from these trials.

#### 2.2.2 Observational studies

We identified three prospective observational studies in this rapid review (Conway Morris et al. 2022, Thuresson et al. 2022, Sloof et al. 2022). Conway Morris et al. (2022) reported on whether HEPA and/or UVC sterilisation had an impact on the amount of SARS-CoV-2 RNA present in various samples during an experiment. Thuresson et al. (2022) reported on the effects of aerosol-generating procedures, HEPA-filtration units, patient viral load, and physical distance. Within this paper, the study reported on the proportion of positive SARS-CoV-2 samples when HEPA filtration was used when compared to no HEPA filtration, along with enhanced verses normal ventilation. Sloof et al. (2022) reported on general particulate matter counts, rather than reporting outcomes relating to SARS-CoV-2 directly.

Conway Morris et al. (2022) conducted an observational study in two repurposed COVID-19 units in the UK where HEPA/UVC sterilisers were installed in a surge ward and an intensive care unit (ICU). Thuresson et al. (2022) conducted an observational study in two hospitals in Sweden whereby some of the outcomes in the paper related to the effectiveness of HEPA filtration units to reduce the presence SARS-CoV-2. The final prospective observational study was published as a pre-print (Sloof et al. 2022). The study aimed to assess aerosol transport within a ward and to determine whether an air filtration unit reduced particulate matter levels and CO_2_ in the air. Using the National Heart, Lung and Blood Institute (NHLBI) Quality Assessment tool for before-after (pre-post) studies, Conway Morris et al. (2022) was deemed to have an overall quality rating of poor whereas Sloof et al. (2022) did not raise many concerns and was deemed to have an overall good quality rating. Using the NHLBI tool for observational cohort and cross-sectional studies, Thuresson et al. (2022) did not raise many concerns and was deemed to have an overall good quality rating.

Conway Morris et al. (2022) aimed to provide evidence for the removal of SARS-CoV-2 and microbial bioaerosols from the air using portable air filters with UVC sterilisation on a COVID-19 ward. In the study, the air filters were placed before the initiation of the three-week study period, switched on at the beginning of week two and run continuously for 24 hours per day. A crossover evaluation was performed where samples were taken to detect SARS-CoV-2 RNA before and after the HEPA/UVC steriliser air filter was introduced in the ward and unit. The study found that when the air filter was continuously running in week two, the samples did not contain any SARS-CoV-2 RNA during any of the five testing days whereas in the ward, during the first week while the air filter was inactive, SARS-CoV-2 was detected on all sampling days. In the ICU, the samples found limited evidence of airborne SARS-CoV-2 in weeks 1 and 3 (when the filter was off) but detected SARS-CoV-2 RNA in a single sample in week 2 (filter on).

Sloof et al. (2022) aimed to determine whether the supplementary air filtration unit was able to reduce particulate matter and CO_2_ levels in the air. The unit contained HEPA filters and an UVC air disinfection lamp (at 254 nm). Particulate matter counts were collected from multiple sensors around the ward over two days (3^rd^ and 4^th^ August 2021). The data was collected from seven automatic sensors every one minute, giving a total of 2782 data points per sensor over the two-day period. However, actual baseline ventilation rates that occurred on the 3^rd^ and 4^th^ August are not reported in the paper as they were unknown. The statistical analysis found that for all the sensors the action of the air filtration unit was associated with a large effect on particulate matters (PM1, PM2.5, CO_2_ and VP levels, which was highly significant. The primary results of the paper found that the air filtration unit greatly reduced particulate matter counts of all sizes throughout the ward space, with a P value of less than 0.001 for all sensors. The particulate matter signals positively correlated with indoor CO_2_ levels with all values reported as statistically significant.

Thuresson et al. (2022) aimed to identify situations, patient characteristics, environmental parameters, and aerosol-generating procedures associated with SARS-CoV-2. Based in two hospitals in Sweden, air samples were collected near patients who were hospitalised with COVID-19 by RT-qPCR. The study collected samples with and without the use of a HEPA filtration unit, as well as samples whereby enhanced ventilation verses normal ventilation was used. Overall, the study favoured the use of HEPA filtration units, stating that by adding a mobile HEPA-filtration unit to rooms with regular ventilation, a reduction in SARS-CoV-2 RNA was found. Compared with rooms with normal ventilation and no HEPA unit (n = 57 samples), rooms with normal ventilation plus an additional mobile HEPA-filtration unit (n = 139 samples) were associated with a reduction in SARS-CoV-2 RNA in the air (odds ratio [OR] 0.3, 95% CI 0.12 to 0.98, p = 0.02). Compared with rooms with normal ventilation (3-4 air changed per hour [ACH]), enhanced ventilation of 8 ACH was associated with a reduction in SARS-CoV-2 RNA in the air (OR 0.3, 95% CI 0.13 to 0.96, p = 0.02).

#### 2.2.3 Modelling and experimental studies

We identified one modelling study and two experimental studies about the inactivation or removal of SARS-CoV-2 using HEPA and/or UVC air filtration systems (Barnewall & Bischoff 2021, Buchan et al. 2020, Snelling et al. 2022).

##### Modelling Study

Buchan et al. (2020) used a high-fidelity coupled radiation-CFD model to quantify the disinfection rates of SARS-CoV-2 within a ventilated room when using far UVC. The experiment distributed SARS-CoV-2 into a private room with and without UVC light. The study found that reductions in SARS-CoV-2 varied depending on which area of the room was sampled and how far away the UVC lamp was from the samples. The study found that the greater number of ACH resulted in quicker rates of viral removal.

Without the UVC lamp, 0.8 ACH ventilation resulted in slow reductions, but quicker reductions when the UVC lamp was used during 0.8 ACH. When the ACH was increased to 8.0 and the UVC lamp was used, the viral removal through ventilation was much quicker (at 45 seconds). Overall, the modelling study reported the combination of far-UVC, and high ventilation rates reduced the viral count the most, which achieved 90% and 99% reductions in approximately 6 and 11.5 minutes, respectively, which was half the time compared to when 8.0 ACH was used during ventilation alone (no UVC). The findings suggest that UVC light combined with 8 ACH can result in disinfection rates of SARS-CoV-2 in a private room, however it is unclear from the literature how many air changes per hour provide the most effective ventilation rates.

##### Experimental Studies

Barnewall & Bischoff (2021) tested the efficacy of an air filtration unit with UVC light and HEPA filtration in a controlled environment using SARS-CoV-2 as the test organism. In the experiment there were three testing groups: the control group, air filtration unit with UVC light, and air filtration unit with UVC light and a HEPA filter. The results of the samples were quantified and found that in the control group where the air filtration unit was not installed, the particulate matter counts were 20. After the introduction of SARS-CoV-2, the particulate matter counts increased to 54. In the UVC group, the particulate matter counts also increased from 69 in the chamber before the introduction of SARS-CoV-2, to 101 in the post-air filtration unit chamber. The most notable decrease in particulate matter counts was observed in the testing group with both UVC light and a HEPA filter, which decreased from 79 to 5.

Snelling et al. (2022) examined the efficacy of a single pass UVC air treatment for the inactivation of coronavirus, MS2 coliphage and Staphylococcus aureus bioaerosols. In the experiment, bovine coronavirus was used as a surrogate for SARS-CoV-2. The model, which was developed for the estimation of the relative risk of exposure to SARS-CoV-2, was adopted to investigate the application of the UVC device to remove COVID-19 RNA in a small medical examination room which measured 15m^2^. The experiment assumed the space had no ventilation with the UVC device implementing 5 ACH with the lowest UVC dose. Of UVC doses 5, 8.27 and 13.27 mJ/cm^2^, the log reduction value was reported at 2.40. The authors note that this result indicated complete inactivation of bovine coronavirus bioaerosols using a single pass UVC air treatment.

#### 2.2.4 Bottom line summary

The limited evidence identified in this review suggests that the use of UVC and/or HEPA filters can reduce viral presence and/or particulates in the air in healthcare settings. In addition, one study suggested that use of HEPA filters did not lead to sleep disturbance. However, we did not identify any evidence directly assessing SARS-CoV-2 transmission or infection rates.

Across all the studies we identified, there was limited reporting and/or variance of many variables that are likely to impact on the results, including room sizes and types, manufacturer or device details, and usability. These factors are important when considering the generalisability, the evidence, and whether the approaches taken in the studies are feasible to implement in current Welsh health care settings and processes.

## 3. DISCUSSION

### 3.1 Summary of the findings

Overall, the evidence identified in this rapid review suggests that UVC light and/or HEPA filtration can reduce particulate matter/viral counts and may also lead to lower symptom scores of COVID-19. Real world evidence suggests supplementary air systems have the potential to reduce SARS-CoV-2 in the air and reduce transmission or infection rates, although further research on how supplementary air filtration systems can reduce transmission is required.

Furthermore, two of the identified papers, the systematic review and one observational study, were assessed as having high risk of bias (Conway Morris et al. 2022, Daga et al. 2021), which should be taken into consideration when drawing conclusions from the outcomes in this review. The systematic review in particular was deemed to have serious limitations with high risk of bias (Daga et al. 2021), which is explored further in Section 3.2.

The type of air filtration systems varied across different studies identified in this rapid review. The systematic review found the use of HEPA filters resulted in fewer symptom scores and less sleep disturbance (Daga et al. 2021), although it is unclear from the paper what the comparative intervention included, and outcomes relating to the transmission or presence of SARS-CoV-2 was not reported in the paper. Two observational studies found that HEPA/UV filtration units reduced the quantity of SARS-CoV-2 observed in testing samples (Conway Morris et al. 2022, Thuresson et al. 2022). Similarly, the pre-print observational study (Sloof et al. 2022) found the HEPA filters and UV light greatly reduced particulate matter counts, although this study did not report on SARS-CoV-2 specifically. The modelling study (Buchan et al. 2020) found similar results, in that the use of UVC light greatly reduced viral concentrations. The study also found that and when combined with eight air changes per hour, viral removal was much quicker when compared with less air changes per hour. The experimental studies (Barnewall & Bischoff 2021, Snelling et al. 2022) also found similar results, in that HEPA/UVC light greatly reduced particulate matter counts.

### 3.2 Critical appraisal

Appendix 1, Table 1 provides an overview of the risk of bias judgements based on a critical appraisal of each included study in this rapid review. Where applicable, we were able to conduct risk of bias evaluations for four papers in this rapid review according to the study design. For the systematic review, the ROBIS tool was used to assess the risk of bias (Whiting et al. 2016). The appraisal evaluates four domains, including the study eligibility criteria, the identification and selection of studies, the data collection and study appraisal, and the synthesis and findings. For two observational studies included in this review (Conway Morris et al. 2022, Sloof et al. 2022), the NHLBI Quality Assessment tool for before-after (pre-post) studies was used (NHLBI 2021) and for Thuresson et al. (2022), the NHLBI Quality Assessment tool for observation cohort and cross-sectional studies was used (NHLBI 2021). We used initial scoping searches to identify suitable tools for critical appraisal of the modelling study. We used a checklist developed by and reported in (Burns et al. 2021). We used the signalling questions listed within this checklist to provide narrative summary which assessed the model structure, input data, methods of validation, how uncertainty was addressed, and transparency of the model/methods. Details of the risk of bias assessment for the modelling study can be found in Appendix 1, Table 2. Risk of bias was assessed independently by two authors. Any differences in appraisal were resolved by consensus.

Caution should be taken when drawing conclusions from both the Daga et al. (2021) systematic review and Conway Morris et al. (2022) due to high risk of bias concerns. For the systematic review (Daga et al. 2021), study eligibility criteria and selection of studies were deemed to be of low concern; however, there were high concerns regarding the data collection and study appraisal, and the synthesis and findings, resulting in overall high risk of bias. Overall, the systematic review mostly reported results as part of a narrative synthesis, with missing details relating to characteristics and outcome data from the studies. Where meta-analysis was performed key detail was also missing, such as which (or how many) studies were included in each pooled estimate. Heterogeneity of results were discussed but not explored, such as through subgroup or sensitivity analyses; authors noted that publication bias was not assessed due to the small number of studies included in each pooled estimate.

Conway Morris et al. (2022) was deemed to be of poor quality. Concerns relate to unclear reporting of aims and objectives, small sample sizes and unclear methodological decisioning making. The additional studies (Sloof et al. 2022, Thuresson et al. 2022) were determined as being of good quality with low risk of bias concerns. The modelling study (Buchan et al. 2020) had low concerns in the risk of bias assessments and was overall deemed to be of good quality. For the modelling study, the input parameters were, to a greater or lesser extent, judged to be transparent, justified, and reasonable. An external validation process was not described; however, the authors reported links to the data for all coding, meaning replication of their methods should be possible.

For the purposes of transparency, the remaining two experimental studies (Barnewall & Bischoff 2021, Snelling et al. 2022) were critically appraised against the modelling critical appraisal checklist. However, we were unable to apply the tool due to the experimental nature of the studies. After further scoping, we were unable to find an appropriate validated risk of bias tool to assess the experimental studies identified in this rapid review. Consequently, there is inherent uncertainty when drawing conclusions from these studies and caution should therefore be taken when applying these results to real world healthcare settings.

### 3.3 Implications for policy and practice

Real world evidence suggests supplementary air cleaning devices have the potential to reduce SARS-CoV-2 presence in the air and transmission rates within healthcare settings, although the evidence base does include studies that were deemed to be of lower priority study designs, and thus, further research is required to develop the evidence on the effectiveness of supplementary air filtration systems.

In general, outcomes relate to the presence of SARS-CoV-2 or general matter counts in samples, in addition to symptom scores and relative risk of exposure. Higher quality evidence and a greater number of studies reporting on the transmission of SARS-CoV-2 in real world healthcare settings are needed to add more certainty to the evidence. Further research to explore the applicability, maintainability, and usability of supplementary air filtration systems in health service settings would be beneficial. It is unknown from the available evidence which type of supplementary filtration system would be the most effective at reducing transmission and/or spread of infection. Further research exploring the effectiveness of different systems, such as different UVC machines, would be beneficial as not all systems are the same and are likely to vary in their effectiveness. Further considerations would also be needed to explore how to optimise the efficacy of the air filtration systems, such as the type of air filtration units and the placement and/or layout of the system in practice, such variables would also impact on the effectiveness of any system used.

Four out of seven papers (Barnewall & Bischoff 2021, Conway Morris et al. 2022, Sloof et al. 2022, Snelling et al. 2022) support the effectiveness of both HEPA filters and UVC light when used together. The studies demonstrate that when HEPA filters and UVC light are used together, a greater reduction in particulate matter counts are observed, although the evidence is based on two observational and two experimental studies. Barnewall & Bischoff (2021) found that the most notable decrease in matter counts was observed in the testing group with both UVC light and a HEPA filter, which decreased from 79 to 5. Two papers explore the effectiveness of HEPA filters alone (Daga et al. 2021, Thuresson et al. 2022) although the systematic review did not report transmission related outcomes. Thuresson et al. (2022) favoured the use of HEPA filtration units, stating that by adding a mobile HEPA-filtration unit to rooms with regular ventilation, a reduction in SARS-CoV-2 RNA was found. Only one modelling study, (Buchan et al. 2020) assessed the use of UVC light, and the findings suggest that UVC light combined with 8 ACH can result in higher disinfection rates of SARS-CoV-2. However, most of the evidence base suggest HEPA filters alongside UV light could provide the most notable reductions in SARS-CoV-2 counts. As the supporting evidence relates to HEPA and/or UVC filtration, the effectiveness of other potential supplementary air filtration systems that could be used remains unclear.

Evidence from the modelling study suggests that quicker viral removal was associated with more air changes per hour, however it is unclear from the evidence what the optimum frequency would be when combined with UVC light and/or HEPA filters. Conway Morris et al. (2022) highlight this research gap, stating that there is a lack of data defining the optimal air changes required to remove detectable pathogens, nor their impact in better ventilated facilities. Buchan et al. (2020) also explored the removal rates of SARS-CoV-2 depending on the area of the sampling room and found that large reductions were seen in the upper regions of a room, whilst small reductions were found where UVC shading was present. Further research in real-life settings exploring where best to place the supplementary air filtration unit would be beneficial.

### 3.4 Strengths and limitations of this Rapid Review

The studies included in this rapid review were identified using a systematic literature review of a range of carefully selected publication databases. The research question and study protocol were developed with significant input from experts in the field. The abstract and full text screening was conducted by two researchers and uncertainty was checked by a third reviewer. The data extraction was performed by a single reviewer and checked for consistency by a second researcher. Whilst the review methods undertaken have been pragmatically robust, there remains the possibility that additional eligible texts have been missed.

This rapid review is limited to few studies that align with the research question and protocol. We are reliant on interpreting the results of studies that have several limitations and this reduces the strength of conclusions. We have made efforts to summarise the key limitations of study designs included in the review and conducted formal risk of bias assessments to assess the quality of the included studies where applicable. It must be noted that whilst this review aimed to extract outcomes relating to SARS-CoV-2 to align with the research question and inclusion criteria, some of the studies that were deemed to be of relevance, list outcomes relating to general particulate matter counts. We have included these outcomes where evidence was limited in relation to the pathogen and setting. As the outcomes discussed in this review varied across studies, this could potentially lead to inconsistent reporting and presentation of findings, thus making it difficult to make clear recommendations.

## Data Availability

All data produced in the present study are available upon reasonable request to the authors

## Abbreviations

Acronym: Full Description
HEPA: High efficiency particulate air
UVC: Ultraviolet C
CFD: Computational Fluid Dynamics
ACH: Air change per hour
VP: Vapour pressure

## 5. RAPID REVIEW METHODS

### 5.1 Eligibility criteria

The aim of this rapid review was to systematically identify and summarise studies that explored the effectiveness of supplementary air cleaning devices in health service settings such as hospitals and dental clinics (including, but not limited to HEPA filtration, UVC light and mobile UVC light devices) to reduce the transmission of SARS-CoV-2.

We have prioritised studies conducted in real world healthcare settings, such as hospitals and dental clinics. However, due to the lack of available data in this context, we have also included modelling and experimental studies that fall within the eligibility criteria outlined in Table 5.

**Table 5:** Inclusion and Exclusion Criteria.

|  | **Inclusion criteria** | **Exclusion criteria** |
| --- | --- | --- |
| **Settings** | Health service settings including, but not limited to hospital settings and dental settings | Non-health service settings such as care homes, classrooms, offices, and restaurants. |
| **Intervention / exposure** | Any supplementary air filtration systems, including (but not limited to): HEPA filtration; UVC light (200-280nm range); Mobile UVC light devices. | UVA/UVB light devices. |
| **Comparison** | Standard air filtration or no air filtration. | Studies comparing different types of supplementary air cleaning devices. |
| **Outcomes** | Any change in transmission related outcomes (compared with baseline or control) for SARS-CoV-2, including change in:<br>▪ presence of virus<br>▪ transmission or infection<br>▪ particulate matter |  |
| **Study design** | We will prioritise the following study types, in the order listed:<br>• Systematic reviews of randomised controlled trials.<br>• Randomised controlled trials.<br>• Non-randomised comparative trials.<br>• Single-arm (no control group) trials that report any relevant outcome.<br>We will prioritise studies conducted in real world settings (i.e., hospital or other healthcare settings).<br>Where data is not available, we will consider experimental studies (lab environment) and modelling studies that align with the research question and eligibility criteria. | Any other modelling studies. |
| **Countries** | Any |  |
| **Language of publication** | English | Any other language |
| **Publication date** | No search date limits applied |  |
| **Publication type** | Published and preprint |  |
| **Reporting of data** | Where possible, we will report on evidence for different types of supplementary filtration systems separately. Where available, we will extract and report on the following variables: specific healthcare setting, room size, temperature, airflow, sampling method, filtration method/device.<br>Where they are reported in evidence relating to SARS-CoV-2, we will report on outcomes for other pathogens. |  |

### 5.2 Literature search

A systematic literature search was conducted between the 20^th^ and 21^st^ July 2022 across a range of databases for English language publications, and then 25^th^ July 2022 for ongoing reviews. No date limit was placed on the search. The search databases and search dates are listed in table 6. Appendix 2 documents the search strategy used for MEDLINE. Search strategies for other databases are available on request.

**Table 6:** Search databases.

| Database | Date Searched |
| --- | --- |
| Ovid MEDLINE | 20<sup>th</sup> July 2022 |
| Ovid Embase | 20<sup>th</sup> July 2022 |
| WHO Global Coronavirus database | 20<sup>th</sup> July 2022 |
| L*VE COVID | 20<sup>th</sup> July 2022 |
| Cochrane C-19 Study Register | 21<sup>st</sup> July 2022 |
| VA-ESP | 20<sup>th</sup> July 2022 |
| Cochrane Library | 20<sup>th</sup> July 2022 |
| Scopus | 21<sup>st</sup> July 2022 |
| PROSPERO | 25<sup>th</sup> July 2022 |

### 5.3 Study selection process and data extraction

Study screening and selection against the eligibility criteria was carried out by Charlotte Bowles, Lauren Elston and Antonia Needham with selection decisions checked by Lauren Elston and any disagreements resolved by consensus amongst the three researchers. Data was extracted as documented in Tables 2-4.

### 5.4 Quality appraisal

Formal quality assessments were completed depending on applicability and study design. Five out of seven studies were formally assessed on their risk of bias. We were unable to find a risk of bias tool that was appropriate for the remaining two experimental studies due to their experimental nature. Further details on the critical appraisal tools used in this rapid review are detailed in section 3.2.

### 5.5 Synthesis

Due to the differing outcome measures and nature of each study, a quantitative analysis of relevant outcomes was not feasible for this rapid review. As a result, evidence was synthesised narratively in relevant sections and individual outcome data are listed in relevant tables within this review.

## 6. EVIDENCE

### 6.1 Study selection flow chart

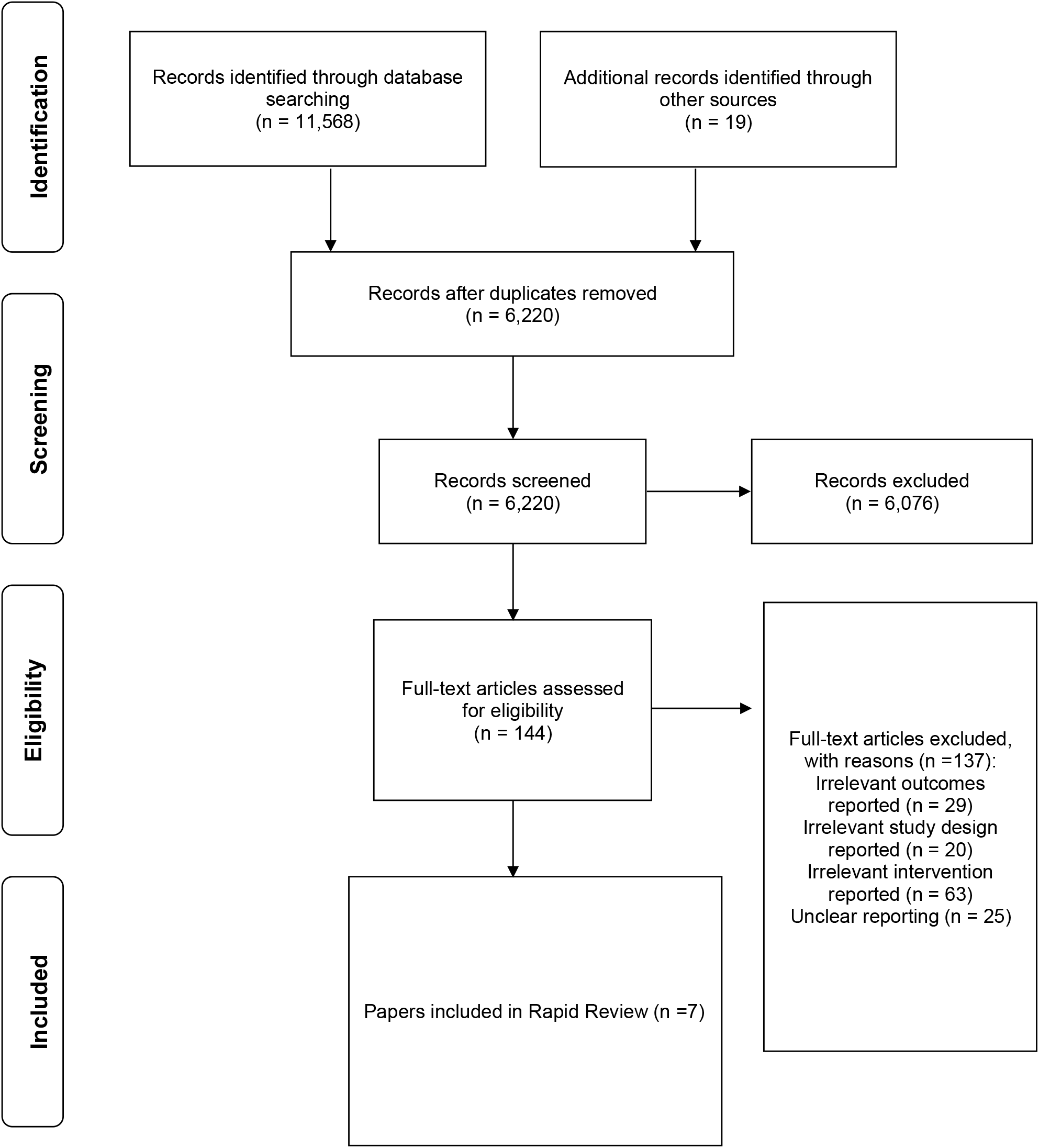

## 7.2 Acknowledgements

The authors would like to thank Davey Jones, Chris Hopkins, Dylan Evans, Geraint Jones, Tony Fisher, Simon Russell, Robert Baker, Catherine Noakes and Robert Hall for their contribution in guiding the focus of the review and to interpreting the findings

## 8. ABOUT THE WALES COVID-19 EVIDENCE CENTRE (WCEC)

The WCEC integrates with worldwide efforts to synthesise and mobilise knowledge from research.

We operate with a core team as part of Health and Care Research Wales, are hosted in the [Wales Centre for Primary and Emergency Care Research (PRIME)](http://www.primecentre.wales/), and are led by [Professor Adrian Edwards of Cardiff University](https://www.cardiff.ac.uk/people/view/123022-edwards-adrian).

The core team of the centre works closely with collaborating partners in [Health Technology Wales](https://www.healthtechnology.wales/), Wales Centre for Evidence-Based Care, [Specialist Unit for Review Evidence centre](https://www.cardiff.ac.uk/specialist-unit-for-review-evidence), [SAIL Databank](https://healthandcareresearchwales.org/about-research-community/secure-anonymised-information-linkage-sail-databank), Bangor Institute for Health & Medical Research/ Health and Care Economics Cymru, and the [Public Health Wales Observatory](https://phw.nhs.wales/services-and-teams/observatory/).

Together we aim to provide around 50 reviews per year, answering the priority questions for policy and practice in Wales as we meet the demands of the pandemic and its impacts.

**Director:**

Professor Adrian Edwards

**Contact Email:**

**Website:**

https://healthandcareresearchwales.org/about-research-community/wales-covid-19-evidence-centre

## 9. APPENDIX 1: Risk of Bias

**Table 1:** Risk of Bias Summary for all included studies.

| Reference | ROB tool used | Overall ROB (concluded based off signalling questions) | Quality Assessment | Comments |
| --- | --- | --- | --- | --- |
| **Daga et al. (2021)** | ROBIS (Whiting et al. 2016) | **HIGH** | **POOR** | Main concerns relate to the synthesis and findings section of the ROB tool. Concerns include: a narrative synthesis of findings, unclear reporting of included studies, heterogeneity not adequately explored, and lack of detail reported for meta-analysis. |
| **Conway Morris et al. (2022)** | NHLBI tool for before-after (pre-post) studies. | **HIGH** | **POOR** | Main concerns relate sample size and sampling methods. |
| **Sloof et al. (2022)** | NHLBI tool for before-after (pre-post) studies. | **LOW** | **GOOD** | Some concerns relating to the generalisability of the population of interest. |
| **Thuresson et al. (2022)** | NHLBI tool for observational cohort and cross-sectional studies. | **LOW** | **GOOD** | Little to no concerns. Some aspects of the tool were not applicable. |
| **Buchan et al. (2020)** | Checklist adapted by (Burns et al. 2021) | **LOW** | **GOOD** | Some concerns relating to external validity. |

**Table 2.**
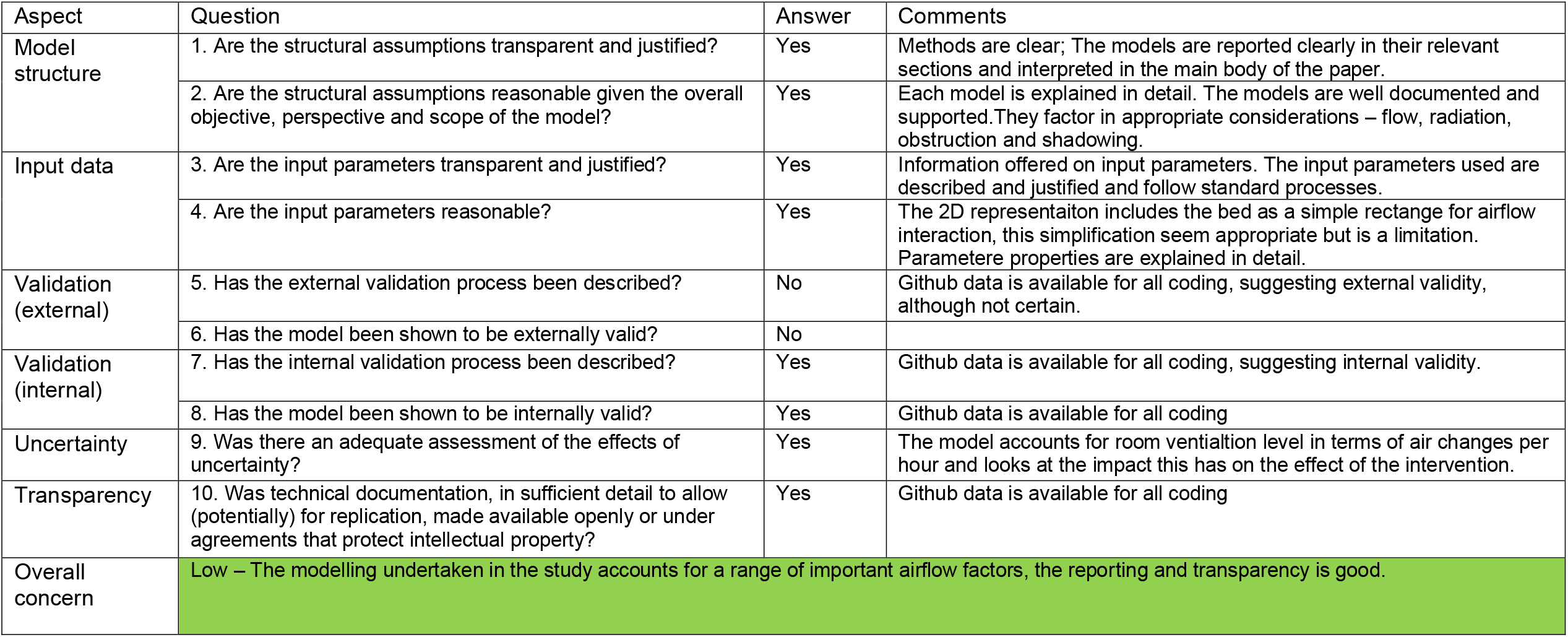
Critical appraisal: Buchan et al. (2020)

## 10. APPENDIX 2: MEDLINE search strategy

10. APPENDIX 2: MEDLINE search strategy Database(s): Ovid MEDLINE(R) ALL 1946 to July 19, 2022
| # | Searches | Results |
| --- | --- | --- |
| 1 | Air Filters/ | 552 |
| 2 | Air Conditioning/ | 2838 |
| 3 | (air filter* or air condition*).tw,kf. | 5133 |
| 4 | (air adj3 (filtrat* or replac* or disinfect* or decontaminat* or clean* or sterili* or saniti* or purif*)).tw,kf. | 6749 |
| 5 | ((filter* or filtrat*) adj (system* or unit*)).tw,kf. | 2710 |
| 6 | (air adj2 scrub*).tw,kf. | 50 |
| 7 | (scrub* adj3 react*).tw,kf. | 30 |
| 8 | Environment, Controlled/ | 4312 |
| 9 | ((engineering or environmental) adj control*).tw,kf. | 3783 |
| 10 | (heat* adj3 ventilat* adj3 air* adj3 condition*).tw,kf. | 376 |
| 11 | HVAC*.tw,kf. | 568 |
| 12 | high* efficien* particulat*.tw,kf. | 514 |
| 13 | (HEPA?? not hepato*).tw,kf. | 2208 |
| 14 | Disinfection/ | 16801 |
| 15 | Decontamination/ | 5548 |
| 16 | Ultraviolet Rays/ | 81847 |
| 17 | (14 or 15) and 16 | 1628 |
| 18 | ((UV* or ultraviolet*) adj3 (germicid* or disinfect* or decontaminat* or inactivat* or clean* or sterili* or saniti* or purif* or scrub*)).tw,kf. | 4796 |
| 19 | ((UV* or ultraviolet* or UV* light or ultraviolet* light*) adj (radiat* or irradiat*)).tw,kf. | 46885 |
| 20 | (ultraviolet and germicidal).tw,kf. | 441 |
| 21 | (ultraviolet-C or ultraviolet C).tw,kf. | 803 |
| 22 | (UVC or UV-C).tw,kf. | 4118 |
| 23 | (computation* fluid* adj3 dynamic*).tw,kf. | 8226 |
| 24 | CFD.tw,kf. | 5901 |
| 25 | *Air Pollution, Indoor/ | 12461 |
| 26 | Air Pollution, Indoor/pc [Prevention & Control] | 2043 |
| 27 | *Air Microbiology/ | 5553 |
| 28 | ((airborn* or air-born*) adj (infection* or pathogen* or transmission*)).tw,kf. | 2313 |
| 29 | Ventilation/ | 6244 |
| 30 | (ventilation adj (system* or unit*)).tw,kf. | 1774 |
| 31 | (building adj3 ventilation).tw,kf. | 221 |
| 32 | or/1-13,17-31 | 108746 |
| 33 | limit 32 to covid-19 | 2265 |
| 34 | limit 33 to english language | 2225 |

## 11. APPENDIX 3: A list of recommended studies that did not meet the inclusion criteria

| Study Citation and Link | Reason for exclusion |
| --- | --- |
| Beggs CB, Sleigh PA. (2002). A quantitative method for evaluating the germicidal effect of upper room UV fields. *Journal of Aerosol Science*. 33(12): 1681-99. doi: [https://doi.org/10.1016/S0021-8502(02)00117-9](https://doi.org/10.1016/S0021-8502%2802%2900117-9) | Outcomes not relevant to SARS-CoV-2. |
| Biasin M, Bianco A, Pareschi G, et al. (2021). UV-C irradiation is highly effective in inactivating SARS-CoV-2 replication. *Scientific Reports* 11(1): 6260. doi: https://doi.org/10.1038/s41598-021-85425-w | Outcomes not relevant to review question. |
| Dreiling JB. (2008). An evaluation of ultraviolet germicidal irradiation (UVGI) technology in health care facilities. Available at: https://hdl.handle.net/2097/651 | Study design and pathogen not relevant to review question. |
| Duan SM, Zhao XS, Wen RF, et al. (2003). Stability of SARS coronavirus in human specimens and environment and its sensitivity to heating and UV irradiation. *Biomedical & Environmental Sciences* 16(3): 246-55. Available at: https://pubmed.ncbi.nlm.nih.gov/14631830/ | Outcomes not relevant to SARS-CoV-2. |
| Jensen MM. (1964). Inactivation of airborne viruses by ultraviolet irradiation. *Applied & Environmental Microbiology*. 12(5): 418-20. doi: https://doi.org/10.1128/am.12.5.418-420.1964 | Outcomes not relevant to SARS-CoV-2. |
| Kowalski W. (2001). Design and optimization of UVGI air disinfection systems. Available at: https://etda.libraries.psu.edu/files/final_submissions/6584 | Study design not prioritised. |
| Kowalski W. (2008). UVGI for hospital applications. *IUVA News*. 10. Available at: https://www.researchgate.net/publication/284449912_UVGI_for_Hospital_Applications | Study design and pathogen not relevant to review question. |
| Kowalski W, Bahnfleth W. (1998). Airborne respiratory diseases and mechanical systems for control of microbes. *HPAC Heating, Piping, Air Conditioning*. 70: 34-48. Available at: https://www.researchgate.net/publication/286967555_Airborne_respiratory_diseases_and_mechanical_systems_for_control_of_microbes | Outcomes not relevant to SARS-CoV-2. |
| Kowalski W, Bahnfleth W. (2000). UVGI design basics for air and surface disinfection. *HPAC Heating, Piping, Air Conditioning*. 72: 100-10 Available at: https://www.researchgate.net/publication/289100700_UVGI_Design_basics_for_air_and_surface_disinfection | Outcomes not relevant to SARS-CoV-2. |
| Kujundzic E, Hernandez M, Miller SL. (2007). Ultraviolet germicidal irradiation inactivation of airborne fungal spores and bacteria in upper-room air and HVAC in-duct configurations. *Journal of Environmental Engineering & Science*. 6(1): 1-9. doi: https://doi.org/10.1139/s06-039 | Outcomes not relevant to SARS-CoV-2. |
| Leach T, Scheir R. (2014). Ultraviolet germicidal irradiation (UVGI) in hospital HVAC decreases ventilator associated pneumonia. ASHRAE Transactions. 120. Available at: https://go.gale.com/ps/i.do?id=GALE%7CA371282999&v=2.1&it=r&linkaccess=fulltext&issn=00012505&p=AONE&sw=w&userGroupName=anon%7Ed4b2415c | Outcomes and study design not relevant to review question. |
| Luo H, Zhong L. (2021). Ultraviolet germicidal irradiation (UVGI) for in-duct airborne bioaerosol disinfection: review and analysis of design factors. Building & Environment. 197: 107852. doi: https://doi.org/10.1016/j.buildenv.2021.107852 | Outcomes and study design not relevant to review question. |
| Nardell EA, Nathavitharana RR. (2020). Airborne spread of SARS-CoV-2 and a potential role for air disinfection. JAMA. 324(2): 141-2. doi: https://doi.org/10.1001/jama.2020.7603 | Study design, intervention and setting not relevant to review question. |
| Noakes CJ, Khan MAI, Gilkeson CA. (2015). Modeling infection risk and energy use of upper-room ultraviolet germicidal irradiation systems in multi-room environments. Science & Technology for the Built Environment. 21(1): 99-111. doi: https://doi.org/10.1080/10789669.2014.983035 | Setting and pathogen not relevant to review question. |
| Reed NG. (2010). The history of ultraviolet germicidal irradiation for air disinfection. Public Health Reports. 125(1): 15-27. doi: https://doi.org/10.1177/003335491012500105 | Study design and outcomes not relevant. |
| Sabino CP, Ball AR, Baptista MS, et al. (2020). Light-based technologies for management of COVID-19 pandemic crisis. Journal of Photochemistry & Photobiology: B, Biology. 212: 111999. doi: https://doi.org/10.1016/j.jphotobiol.2020.111999<br>https://doi.org/10.1016/j.jphotobiol.2020.111999 | Study design, intervention, and outcomes not prioritised. |
| Saran S, Gurjar M, Baronia A, et al. (2020). Heating, ventilation and air conditioning (HVAC) in intensive care unit. Critical Care. 24: 194. doi: https://doi.org/10.1186/s13054-020-02907-5 | Intervention and study design not relevant to review question and outcomes. |
| Sodiq A, Khan MA, Naas M, et al. (2021). Addressing COVID-19 contagion through the HVAC systems by reviewing indoor airborne nature of infectious microbes: will an innovative air recirculation concept provide a practical solution? Environmental Research. 199: 111329. doi: https://doi.org/10.1016/j.envres.2021.111329 | Setting and intervention not relevant to review question. |
| Walker CM, Ko G. (2007). Effect of ultraviolet germicidal irradiation on viral aerosols. Environmental Science & Technology. 41(15): 5460-5. doi: https://doi.org/10.1021/es070056u | Study design, pathogen, and outcomes not relevant to review question. |

